# Bridging the gaps in test interpretation of SARS-CoV-2 through Bayesian network modelling

**DOI:** 10.1101/2020.11.30.20241232

**Authors:** Yue Wu, David Foley, Jessica Ramsay, Owen Woodberry, Steven Mascaro, Ann E Nicholson, Tom Snelling

**Affiliations:** School of Public Health, The University of Sydney, Camperdown, New South Wales, Australia; Department of Infectious Diseases, Perth Children’s Hospital, Perth, Western Australia, Australia; Wesfarmers Centre for Vaccines and Infectious Diseases, Telethon Kids Institute, Perth, Western Australia, Australia; Department of Data Science & Artificial Intelligence, Monash University, Clayton, Victoria, Australia; School of Public Health, Curtin University, Bentley, Western Australia, Australia; Menzies School of Health Research, Charles Darwin University, Darwin, Northern Territory Australia

**Keywords:** SARS-CoV-2, RT-PCR test, diagnostic decision support, false negative, causal diagram, Bayesian belief model

## Abstract

**Background:** In the absence of an established gold standard, an understanding of the testing cycle from individual exposure to test outcome report is required to guide the correct interpretation of SARS-CoV-2 reverse transcriptase real-time polymerase chain reaction (RT-PCR) results and optimise the testing processes. Bayesian network (BN) models have been used within healthcare to bring clarity to complex problems. We use this modelling approach to construct a comprehensive framework for understanding the real world predictive value of individual RT-PCR results.

**Methods:** We elicited knowledge from domain experts to describe the test process from viral exposure to interpretation of the laboratory test, through a facilitated group workshop. A preliminary model was derived based on the elicited knowledge, then subsequently refined, parameterised and validated with a second workshop and one-on-one discussions.

**Results:** Causal relationships elicited describe the interactions of multiple variables and their impact on a RT-PCR result. Some interactions are infrequently observable and accounted for across the testing cycle such as pre-testing factors, sample collector experience and RT-PCR platform. By setting the input variables as ‘evidence’ for a given subject and preliminary parameterisation, three scenarios were simulated to demonstrate potential uses of the model.

**Conclusions:** The core value of this model is a deep understanding of the total testing cycle, bridging the gap between a person’s true infection status and their test outcome. This model can be adapted to different settings, testing modalities and pathogens, adding much needed nuance to the interpretations of results.

## Introduction

Effective containment of COVID-19 disease caused by the severe acute respiratory syndrome coronavirus 2 (SARS-CoV-2) rests upon the rapid and accurate identification of cases. Although nucleic acid amplification tests, including real time reverse transcriptase polymerase chain reaction (RT-PCR), are widely used, the absence of an established gold standard diagnostic method has hindered the assessment of test performance ^1^. The potential for false negative results is well-recognised; such results can significantly undermine the public health response, facilitating ongoing chains of transmission. Similarly, at the patient level, it may delay case recognition, place other patients and healthcare workers at risk and, importantly, impede the commencement of emerging treatments. A wide variation in rates of false negatives has been reported, ranging from 1.8 to 58% ^2^; this variability may be attributable to heterogeneity in disease prevalence, patient age, timing of testing, type of specimen, other components of the pre-analytical phase and the RT-PCR assay employed across studies ^3^.

Although better standardisation of data collection and reporting may add further clarity, a comprehensive understanding of the mechanisms involved in testing is required to help develop strategies to improve testing systems and importantly, guide the correct interpretation of test results within the associated context. This includes a true understanding of the positive and negative predictive values of a test at a national, regional and patient level and the potential to permit the early identification of false negative results. The limitations in available data undermine these efforts. An alternative method is required to bridge the current gaps in knowledge.

Bayesian network (BN) modelling offers an approach to understanding complex problem domains by organising information, whether directly observable or not (i.e. latent), under a causal inference framework ^4,5^. A BN is a probabilistic graph model that integrates available data with subject-matter knowledge from domain experts to describe how a system operates ^6^. BNs have been used within healthcare to improve clinical decision-making^7^, bringing clarity to complex problems, especially where there is little quality data available^7^.

We elicited knowledge from a range of domain experts to construct a causal BN which describes the testing process for SARS-CoV-2 by RT-PCR, from individual exposure through to the interpretation of the laboratory test result. In explicitly modelling the latent trajectory of a pathogen through its diagnostic pathway, we have generated a common framework which accounts for a range of factors that plausibly influence test results, and which may be generalizable to other pathogens and assay formats.

## Methods

A BN comprises two parts: 1) a graph that uses nodes to represent the factors (or variables) which are relevant to describing and understanding the system, and arrows to represent the direct statistical (and often causal) dependencies between them; and 2) a set of conditional probability distributions that specifies the strength of each of those dependencies, which then forms a joint probability distribution over all variables.

Clinical experts in microbiology, infectious diseases and general medicine contributed their relevant subject matter knowledge. Key variables were identified through literature review and an online discussion with the experts. Knowledge elicitation was guided by trained facilitators, defined as knowledge engineers, and supported utilising graphical representations of interactions between variables within the proposed structure ^6,8^. A preliminary model structure was created via a subsequent group knowledge elicitation workshop; this was then reviewed and refined in one-on-one discussions with the experts. A preliminary parameterisation of the model was performed to produce qualitative behaviour that matched the modellers’ and the experts’ high level understanding of the problem domain. The refined model structure was reviewed and validated by experts in a second workshop and one-on-one discussions.

We provide a narrative description of the model structure and illustrate its potential application in three scenarios. All nodes are labelled and referred to by numbers ***1-31***. The term ‘virus’ and all described events relate to SARS-CoV-2, unless stated otherwise. The model was built in GeNIe (https://www.bayesfusion.com/downloads/). Appendix A provides a comprehensive variable dictionary for this model, with references to justify each node and arc where possible. Detailed conditional probability tables can be accessed via ‘testing_v7.6_params.xdsl’ on the OSF platform, which will also include any future updates.^i^

## Results

Figure 1 shows the simplest possible BN for representing true and false positive and negative rates produced by laboratory results.^ii^ The figure illustrates this simple BN in two scenarios: when a person is infected now (left) and when a person is not infected now (right). According to this BN, there is an 85% chance of detecting the virus if a person is infected at the time of testing, giving a corresponding false negative probability of 15%; and a 0.1% chance of falsely detecting the virus if a person is not infected, giving a corresponding 99.9% probability of a true negative. Obtaining accurate estimates of these rates is challenging because we cannot directly observe (nor perfectly control) the true infection status at the time of testing – the very reason a test is needed – and hence must make do with general estimates based on controlled samples. We can, however, make improved case-specific estimates by incorporating factors involved in the process of sampling and testing into our model. We can also improve our assessment of whether a person is truly infected by incorporating background factors (such as age) that may influence both the prior probability of an infection as well as other factors related to the testing process (such as the chance of finding virus at a particular sample site). The model we present next describes how we have expanded this simple model to include other relevant variables that interact to drive changes to sensitivity, specificity and, ultimately, the probability of infection.

**Figure 1.**
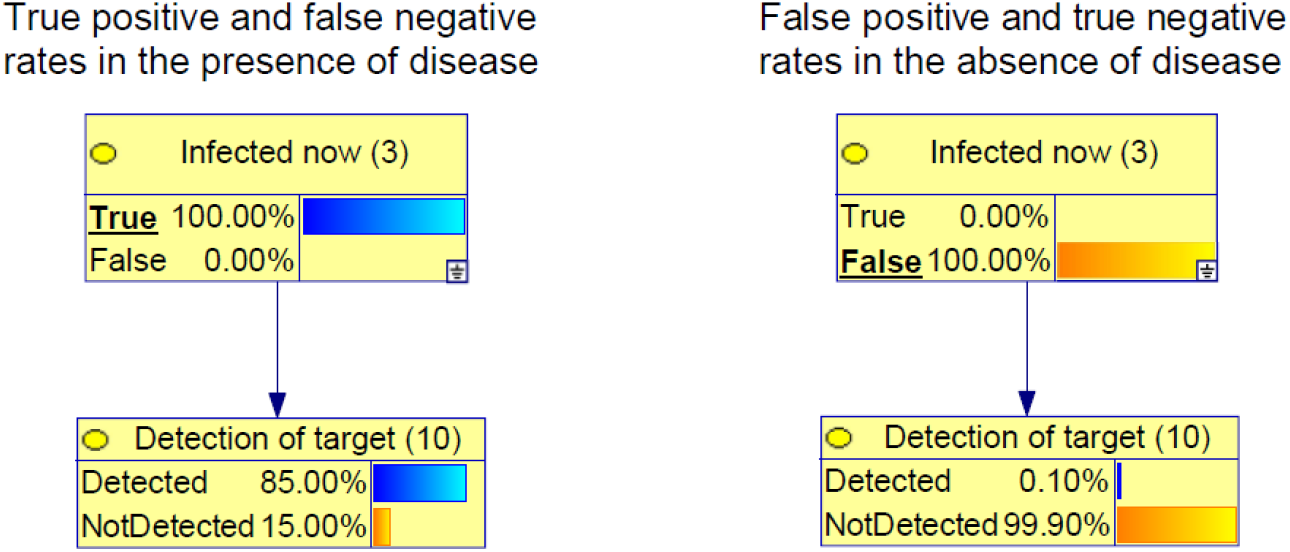
Definitions of true and false positives and negatives for laboratory results. (Left) The true positive rate is the probability of Detected amongst those infected and the false negative rate is the probability of NotDetected also amongst those infected. (Right) The false positive rate is the probability of Detected amongst those who are **not** infected and the true negative rate is the probability of NotDetected also amongst those who are not infected.

### Model description

The expanded BN (Figure 2) models the trajectory of SARS-CoV-2 as it’s sampled, transported, extracted and amplified, along with the conditions and operations that can affect the sample throughout this process.^iii^ The SARS-CoV-2 trajectory itself is modelled via a sequence of latent nodes (coloured yellow, ***1-10***), running down the centre of the graph, with the previously introduced **infected now** (***3***) and **detection of target** (***10***) sitting at almost opposite ends of this sequence. The probability of being **infected by a known viral exposure** (***2***) is driven by the **intensity of that exposure** (***1***). If infected by the known exposure (***2***), **age** (***11***) and the number of **days since the exposure** (***12***) influence the probability of being **infected at the time of testing** (***3***), and also drive the **days since first compatible symptom onset** (***13***). The probability of infection from an unknown exposure is possible and currently parameterised to be low, although this risk is influenced by the background prevalence of the virus in a given population at a given time, and therefore needs to be calibrated according to the setting. The background probability of compatible symptoms unrelated to a known exposure (potentially non-SARS-CoV-2) is also set to be low, and as a result, the presence of symptoms predicts a high probability of being infected by a recent exposure. However, this background probability is also driven by the circulation of other symptom-compatible pathogens.

**Figure 2.**
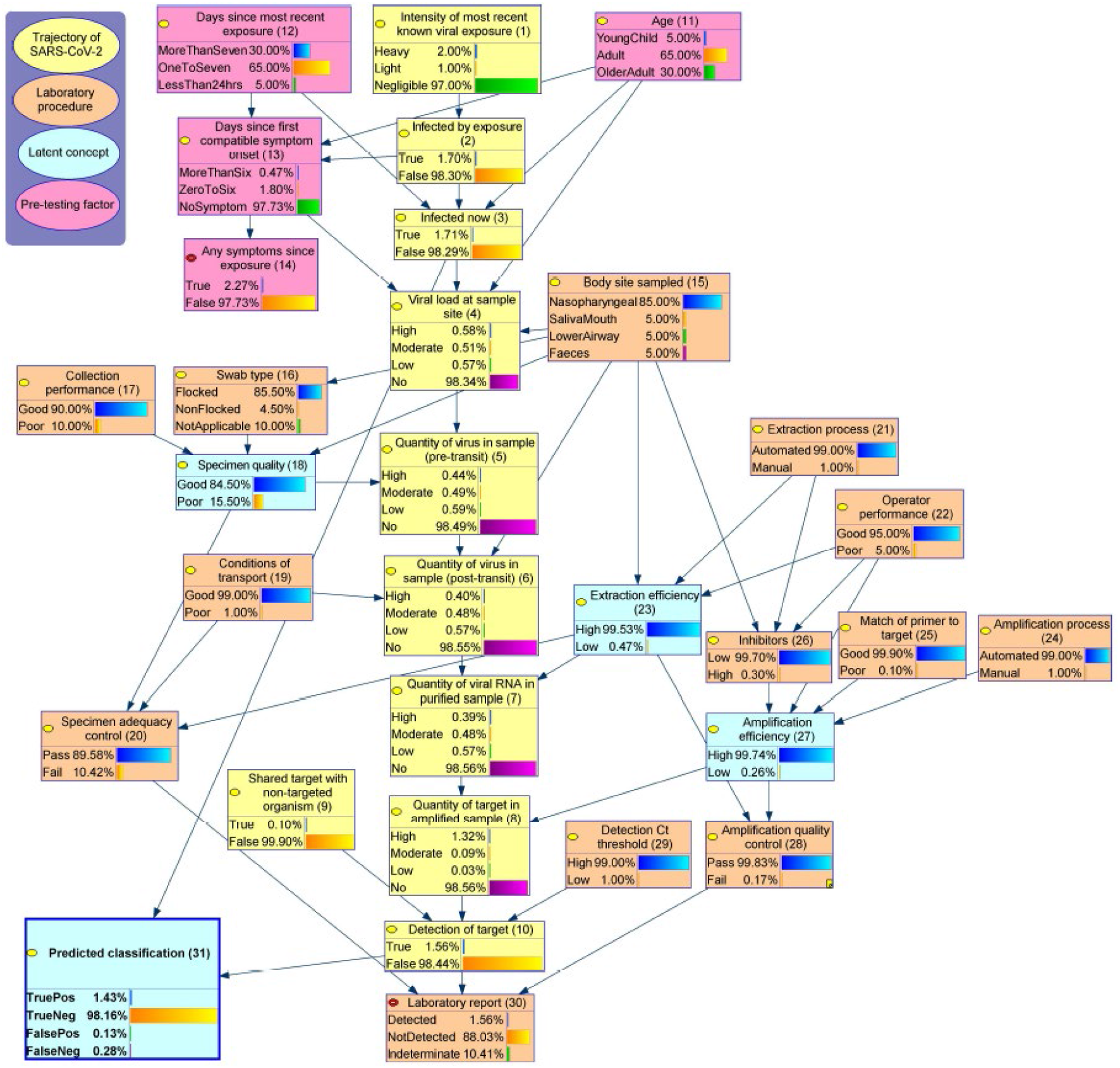
The causal Bayesian network of RT-PCR testing of SARS-CoV-2. This diagram presents the model structure, variable values, and marginal distributions (i.e. when nothing is known, other than that a test has been conducted). Appendix A provides a comprehensive variable dictionary for this model. Detailed conditional probability tables can be accessed ‘testing_v7.6_params.xdsl’ on the OSF platform.

Among those with **SARS-CoV-2 infection at the time of testing** (***3***), the **viral load at a given sample site** (***4***) is influenced by the number of **days since first symptom onset** (***13***)^iv^, **body site sampled** (***15***) and **age** (***11***). In particular, the model assumes the viral load in the upper airway is initially highest, followed by increasing amounts in lower respiratory tract and faeces over time.

When collecting a sample, the quantity of virus obtained from the **viral load at sample site** (***4***), equating to the **quantity captured in sample (pre-transit)** (***5***) depends on the **specimen quality** (***18***), a latent variable which captures the technical and operator-dependent factors which affect the adequacy of collection. **Specimen quality** (***18***) is therefore improved by a good **collection performance** (***17***) (e.g., indicated by collector’s experience), the use of a flocked **type of swab** (***16***) (if applicable) and a site that requires a simpler collection technique, such as **specimen sampled from** Saliva and Mouth sites (***15***). The **quantity of virus in sample post-transit** (***6***) may be affected by **the quantity of virus in the sample pre-transit** (***5***), the **conditions of transport** (***19***) and **body site sampled** (***15***); for example, faecal specimens may contain substances which accelerate the degradation of viral nucleic acid.

In the laboratory, the **extraction** and **amplification processes** (***21, 24***) are assumed to be predominantly automated, meaning a testing process that is less affected by **operator performance** (***22***), compared with manual methods. In addition, a high level of **inhibitors** (***26***) and a poor **match of primer to target** (in the virus) (***25***) can reduce **amplification efficiency** (***27***). Low **extraction** and **amplification efficiency** (***23, 27***) (both latent) may increase the probability of a false negative results if the **quantity of virus** is low in the **post-transit** (***6***) and **purified samples** (***7***), respectively. The probability of a false negative result may also increase if the **detection Ct threshold** (***29***) is lowered (e.g., to 35). A false positive result may occur if the specimen contains a **shared target from a non-SARS-CoV-2 organism** (***9***). Similarly, if the **detection Ct threshold** (***29***) is set higher (e.g., 40 and above), the risk of non-specific amplification increases and, consequently, the risk of a false positive result. These events are assumed to be rare by the current model.

Finally, the **lab report** (***30***) will be detected if the **viral target is detected** (***10***), often described as a positive result. If the target is not detected, the test result will be reported as not detected if the specimen has passed both the **specimen** and **amplification quality controls** (***20, 28***) (a negative result), and as indeterminate otherwise (where a repeat test may be requested). In cases where the **SARS-CoV-2 target** (***10***) is *not detected*, there is a high likelihood that this represents a true or false negative result if the probability of being **infected now** (***3***) is low or high, respectively, and likewise for true or false positive. This relationship is now described using the node **predicted classification** (***31***).

### Example scenarios

Three illustrative scenarios were developed in conjunction with the experts. The model outputs were obtained by setting the input variables as ‘evidence’ for a given constructed scenario. Please access appendix B for detailed models with input variables selected for each scenario.

#### *Scenario 1:* The predicted probability of infection and testing results influenced by exposure intensity and presence of symptoms

Consider an **older adult** (***11***) who had a **light exposure** (***1***) to the virus **1 to 7 days ago** (***12***) (e.g. brief contact in a cafe) but with **no symptoms** (***13***) currently. The probability of this person **currently being infected** (***3***) is estimated to be 2.3%, and the probability of returning a **positive nasopharyngeal swab** (***15***) result is 1.9% (***30***) with predicted 0.5% chance being a **false negative** (***31***). However, if the **intensity of exposure was heavy** (***1***) (e.g., household contact), the risk of **being infected** (***3***) would be 44.2% and the probability of returning a **positive test result** (***30***) would be 35.3% with **false negative prediction** increased to 9.0% (***31***). Rather than having no symptoms, if the person experienced onset of **symptoms 0 to 6 days** (***13***) **after that exposure** (***1***), the probability of **being infected** (***3***) is estimated to be 98.7%, and the probability of returning a **positive result** (***30***) is 94.5% (**false negative** (***31***) 4.3%).

#### *Scenario 2:* Influence of specimen quality on the probability of a positive test result in those who are infected

Consider the same **older adult** (***11***) who was **heavily exposed** (***1***) **to the virus 1 to 7 days ago** (***12***) and had onset of **symptoms 0 to 6 days afterwards** (***13***). Consider now that this person **is infected** (***3***). A **nasopharyngeal swab** (***15***) is taken for testing. If a **poor collection** (***17***) is performed with a **non-flocked swab** (***16***) and the **conditions of transport** (***19***) are poor, the probability that the lab reports a **positive result** (***30***) is 89.2%, the probability of an indeterminate result is 6.5%, and the probability of a false negative result (***31***) is 10.8%. However, for a **good collection performance** (***17***) using a **flocked swab** (***16***) and with **good specimen transport conditions** (***19***), the probability of a **positive result** (***30***) is 95.9%, the probability of an indeterminate result is 0.6%, and the probability of a false negative result (***31***) is 4.1%.

#### *Scenario 3:* Understanding the patient characteristics of those with false negative test results

Individuals who are truly **infected at the time of testing** (***3***) but who **tested negative** (***30***) (i.e. false negatives) are **younger** (***11***) and have a higher probability (63.7%) of having **no symptom at the time of testing** (***13***) than those who are infected and **test positive** (***30***) (22.4%).

## Discussion

Accurate diagnosis of COVID-19 is critical to guide patient management, including infection control and public health responses^9^. Although there is increasing data on the performance of commercial assays ^10^, these assessments typically use non-clinical samples and are performed in closely controlled environments. A range of variables, from the age of the subject, the nature of exposure, the presence and duration of symptoms, operator skill and assay technical complexity can all influence the positive and negative predictive value of a test and are therefore important considerations when interpreting any test result. The core value of this model is its explicit representation of these variables and their probabilistic interdependencies, allowing a deeper understanding of test results by explicitly defining true positive, true negative, false positive, and false negative interpretations based on the discrepancy between a person’s true infection status and their test outcome. On a population level, the model can demonstrate how and where improvements in processes and procedures may improve the value of the test. Given a fully parameterised model, tracking changes in the distribution of these variables (depicted in scenario three) over time and across settings can help understand how public health responses can be optimised for the timely detection of cases^9^ so that effective containment strategies can be implemented.

When the model is applied to a single patient, it can also inform individual-level management. The probability that a person is infected can be more correctly inferred by integrating the test result with knowledge of the background risk (or ‘pre-test probability’), the intrinsic assay characteristics, and the adequacy of the sampling and laboratory procedures. This is illustrated in scenario one where there is still a 4.3% chance that a false negative result will be obtained in an older patient with infection and symptoms, which could have significant ramifications in terms of that patient’s outcome, and the risk of spread. Importantly, if a negative result was obtained, the model would allow this result to be reviewed in context, guiding the clinician’s interpretation of the result through a better understanding of the negative predictive value for that individual patient.

Causal BNs allow the exploration and characterisation of a complex problem based on elicited knowledge from domain experts, even when limited data is available; a valuable characteristic during an outbreak of a novel pathogen. The model allows inclusion of known components of the testing cycle, including specimen collection and transport^11,12^, elements that are often not known when interpreting the result. Specimen adequacy can influence the amount of virus present at the site that is “collected” for testing. Poor collection performance may reduce the advantage of the more technically difficult to collect specimen^13^. Scenario two underlines the importance of a good specimen collection, coupled with other factors, decreasing the probability of a false negative from 10.9% to 4.1%. Similarly, although mouth and saliva swabs are technically easier to collect, better tolerated and may facilitate self-collection, they have a potentially lower predictive value due to the lower quantity of virus at that site ^13^.

The expanding drive for testing and pressure of rapid turnaround times places enormous strain on laboratory staff and testing systems. Although capacity has increased, the impact of the human element can’t be underplayed^14^ and needs to be accounted for when considering the predictive value of a result; inexperienced staff, extended work hours and increased pressure can impact the numerous intricate steps of laboratory testing and thereby affect test performance. Automated processes can mitigate some potential errors, yet are not available in all settings across the globe. Laboratory quality assurance measures including extraction controls may help to identify systematic errors and reduce false negatives secondary to poor extraction or the presence of inhibitors.

COVID-19 RT PCR assays have been designed to match the novel emerged virus. A future concern, included in the model, is the potential drift and divergence of COVID-19 strains into distinct lineages. These changes may alter the amplification site, reducing the RT-PCR ability to detect the presence of different lineages. Similar evolutionary changes have been observed in influenza, requiring re-tooling of the nucleic acid amplification^15^.

The model can be calibrated to account for the changing population incidence of COVID-19 and adjusted for low or high viral incidence rates. Rates of co-circulating pathogens can also be incorporated into the model. For example, respiratory syncytial virus and influenza were low in Australia^16^ during the winter of 2020; in the model this would increase the probability that respiratory symptoms after an exposure would be suggestive of COVID-19 infection. As the northern hemisphere enters their winter, the model can be calibrated to reflect their rates over their typical peak season. The model also has the flexibility to be modified based on application to account for the different performance of RT-PCR platforms used in different laboratories, as well as for other pathogens.

## Limitations

To guide individual and public health decision-making, the model will need to be validated using data. Expert opinion may incorrectly guide the model, as current knowledge and experiences may not be generalizable to this outbreak. Further setting-specific parameterisation and validation of the model is required before introducing into a real-world setting. These should involve important parameters such as location and population specific prevalence of SARS-CoV-2 and other respiratory viruses with compatible symptoms, and the changing viral load at each sample site at the different time points post infection.

## Conclusion

Bayesian networks have the potential to improve decision-making and clinical care, especially around the correct interpretation of a COVID-19 clinical diagnostic test, such as RT-PCR. This model could be further strengthened by expanding it to include other nucleic acid amplification platforms and other testing modalities.

## Data Availability

Detailed conditional probability tables can be accessed via testing_v7.6_params.xdsl on the OSF platform, which will also include any future updates.

https://osf.io/x5c4u/?view_only=afbdfb3e22c2406cad1ae142a3e5a3b2

## Acknowledgement

We are grateful to the following experts who participated in the knowledge elicitation sessions: Mark Nicol, Susan Benson, Ben Scalley, Edward Raby, Jen Kok, Matthew O’Sullivan, Ariel Mace, Charlie McLeod, Christopher Blyth, Gladymar Perez, Mark Boyd, Mejbah Bhuiyan. Funding for this project has been provided by the Digital Health Cooperative Research Centre and the Snow Medical Research Foundation.

---

**Appendix A.** Variable dictionary

| Variable name | Description | Potential values | Status | Parent nodes | References |
| --- | --- | --- | --- | --- | --- |
| **Intensity of most recent known viral exposure (1)** | Intensity of most recent exposure to SARS-CoV-2. This variable can be influenced by many epidemiological factors not covered in this current model. | *Heavy,*<br>*Light,*<br>*Negligible* | Latent | None | 17 |
| **Infected by exposure (2)** | Whether the person was infected due to the most recent exposure. | *True,*<br>*False* | Latent | Intensity of most recent known viral exposure |  |
| **Age (11)** | Age of person tested. | *YoungChild,*<br>*Adult,*<br>*OlderAdult* | Observable | None | 18,19 |
| **Days since most recent exposure (12)** | On the day of testing, the number of days since most recent known exposure to SARS-CoV-2. | *MoreThanSeven,*<br>*OneToSeven,*<br>*LessThan24hrs* | Observable | None | 20-22 |
| **Days since first compatible symptom onset (13)** | On the day of testing, the number of days since the onset of first (compatible) symptom. It is unknown whether the intensity of exposure influences the timing of symptom onset, so no corresponding arc has been included. | *MoreThanSix,*<br>*ZeroToSix,*<br>*NoSymptom* | Observable | Days since most recent exposure, age, infected by exposure | 12,20,22,23 |
| **Any symptoms since exposure (14)** | On the day of testing, whether there have been any symptoms since the most recent exposure. This (deterministic) node was created for informational purposes. | *True,*<br>*False* | Observable | Days since first symptom onset | 23 |
| **Infected now (3)** | SARS-CoV-2 infection (at any body site) at the time of testing. If not infected by the most recent known exposure, the risk of being infected now is currently set to be 0.1%. This probability should be driven by the spatial and temporal prevalence of the virus in a given setting. | *True,*<br>*False* | Latent | Days since most recent exposure, infected by exposure, age |  |
| **Body site sampled (15)** | Body site swabbed or bodily fluid collected for SARS-CoV-2 testing. A non-uniform prior is used for this node, with the majority (85%) sample site set to nasopharynx. | *Nasopharyngeal,*<br>*SalivaMouth,*<br>*LowerAirway,*<br>*Faeces* | Observable | None | 12,24,25 |
| **Viral load at sample site (4)** | Amount of SARS-CoV-2 at sample site. The CPT of this node may vary by site which interacts with the progression of disease. | *High,*<br>*Moderate,*<br>*Low,* | Latent | Infected now, days since first compatible | 26-29 |
|  |  | *No* |  | symptom onset, age,<br>body site sampled |  |
| **Swab type (16)** | Type of swab used to obtain SARS-CoV-2 sample. This variable is only applicable to the sites sampled by swab. | *Flocked,*<br>*NonFlocked,*<br>*NotApplicable* | Observable | Body site sampled | <sup>30, 31</sup> |
| **Collection performance (17)** | The resulting performance of the sample collection process. This may be influenced by various factors (not explicated), including the ability of the person collecting the sample for SARS-CoV-2 testing. | *Good,*<br>*Poor* | Observable | None | <sup>32,33</sup> |
| **Specimen quality (18)** | This is a latent concept that summarises the quality of the specimen, collected for SARS-CoV-2 testing, in terms of its ability to confer an accurate result. | *Good,*<br>*Poor* | Latent | Collection performance,<br>swab type, body site<br>sampled | <sup>12,21,24,34-38</sup> |
| **Quantity of virus in sample (pre-transit) (5)** | The number of viral particles obtained for testing within the sample. | *High,*<br>*Moderate,*<br>*Low,*<br>*No* | Latent | Viral load at sample site,<br>specimen quality |  |
| **Conditions of transport (19)** | The conditions the specimen is subjected to, from collection to laboratory processing. This includes temperature, transport time, transport with/without viral transport media, which are observable but not explicitly described in the current model. | *Good,*<br>*Poor* | Observable | None | <sup>11,12</sup> |
| **Quantity of viable virus in sample (post-transit) (6)** | The remaining amount of detectable virus in the sample after exposure to transport conditions. | *High,*<br>*Moderate,*<br>*Low,*<br>*No* | Latent | Conditions of transport,<br>body site sampled,<br>quantity of virus in<br>sample pre-transit |  |
| **Extraction process (21)** | The process by which the nucleic acid is extracted from the collected sample. Extraction process can differ with manual and automated processes. Prior distribution sets 99% to be automated, however this might not be the case in resource-poor settings. | *Automated,*<br>*Manual* | Observable | None | <sup>12,39</sup> |
| **Operator performance (22)** | The accuracy of the scientist/technician performing the PCR testing procedures in the lab. In addition to experience, this may also be influenced by other factors, such as workload, which are not covered in the current model. | *Good,*<br>*Poor* | Observable | None | <sup>14,40</sup> |
| **Extraction efficiency (23)** | The collective quality of the nucleic acid extraction process to produce a sample able to detected SARS-CoV-2 if present. | *High,*<br>*Low* | Latent | Body site sampled, extraction process, operator performance | 41,42 |
| **Quantity of viral RNA in purified sample (7)** | The amount of SARS-CoV-2 viral RNA extracted from the sample. | *High,*<br>*Moderate,*<br>*Low,*<br>*No* | Latent | Quantity of virus in sample post-transit, extraction quality, operator performance | 12 |
| **Inhibitors (26)** | Any factor which prevents the amplification of SARS-CoV-2 RNA during the PCR testing of the sample. | *Low,*<br>*High* | Latent | Conditions of transport, body site sampled, extraction process, operator performance | 12,43,44 |
| **Amplification process (24)** | The process of replicating the SARS-CoV-2 RNA sequence(s) millions of times. Prior distribution sets 99% to be automated, however this might not be the case in resource-poor settings. | *Automated,*<br>*Manual* | Observable | None | 41 |
| **Match of primer to target (25)** | The closeness in similarity of the nucleic acid sequence used in the PCR assay to the nucleic acid sequences in the extracted RNA from patient sample. Prior distribution sets 99.9% to be good match, especially as there is often more than one target selected for the primer. | *Good,*<br>*Poor* | Observable | None | 45-48 |
| **Amplification efficiency (27)** | The collective quality of the test sample after the amplification process. | *High,*<br>*Low* | Latent | Inhibitors, operator performance, match of primer to target, amplification process |  |
| **Quantity of target in amplified sample (8)** | The amount of RNA in the sample present after amplification that specifically matches the primers and probes of the PCR testing kit (summarised by amplification efficiency). | *High,*<br>*Moderate,*<br>*Low,*<br>*No* | Latent | Quantity of viral RNA in purified sample, amplification efficiency |  |
| **Specimen adequacy control (20)** | A Sample Adequacy Control (SAC) targets a single copy human gene that should be present in each specimen. The SAC confirms adequate patient sample has been collected and appropriate testing has occurred. | *Pass,*<br>*Fail* | Observable | Specimen quality, conditions of transport, extraction efficiency | 44 |
| **Amplification quality control (28)** | A nontarget nucleic acid sequence coamplified with the sample to measure the quality of the amplification process. | *Pass,*<br>*Fail* | Observable | Amplification efficiency, extraction efficiency | 47,49 |
| **Detection Ct threshold (29)** | The number of cycles set within a PCR system determines when a fluorescent signal exceeds the background level indicating a positive sample. A higher threshold allows greater amplification, in turn allowing the system to be more sensitive. A low threshold may be 35 and a high threshold may be 40. The prior has been set to a 99% chance of using a high (and therefore more sensitive) Ct threshold. | *High,*<br>*Low* | Observable | None | 20,26,49 |
| **Shared target with non-targeted organism (9)** | The specificity of the SARS-CoV-2 nucleic acid sequence within the PCR assay to the SARS-CoV-2 within the sample tested. Prior distribution sets 99.9% to be false, however, if true, this (and this alone) introduces the risk of false positives. | *True,*<br>*False* | Observable | None | 46,47 |
| **Detection of target (10)** | Detection of the nucleic acid sequence specific to the SARS-CoV-2 virus targeted by the PCR assay. | *True,*<br>*False* | Observable | Shared target with non-targeted organism, quantity of target in amplified sample, detection Ct threshold | 50 |
| **Laboratory report (30)** | The reported result of the SARS-CoV-2 PCR assay. This is set to be a deterministic node that is Detected if detection of target was true; or, in the absence of detection, NotDetected if quality controls were passed and Indeterminate otherwise. | *Detected,*<br>*NotDetected,*<br>*Indeterminate* | Observable | Detection of target, amplification control, specimen quality control | 51,52 |
| **Predicted classification (31)** | Model prediction expressing the predictive probability of a true positive, true negative, false positive, and false negative | *TruePos,*<br>*TrueNeg,*<br>*FalsePos*<br>*FalseNeg* | Latent | Viral load at sample site, detection of target |  |

**Appendix B.**
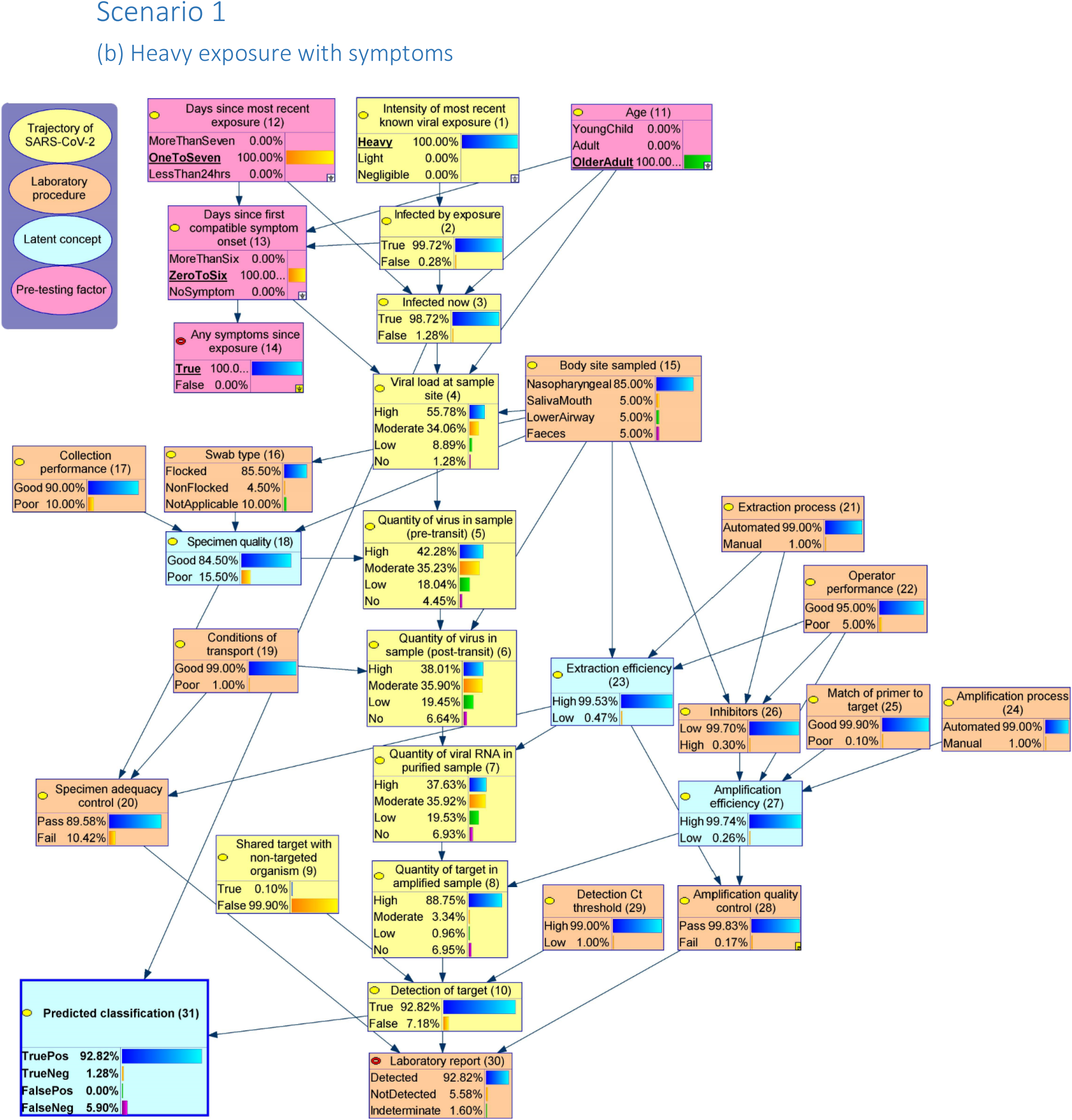
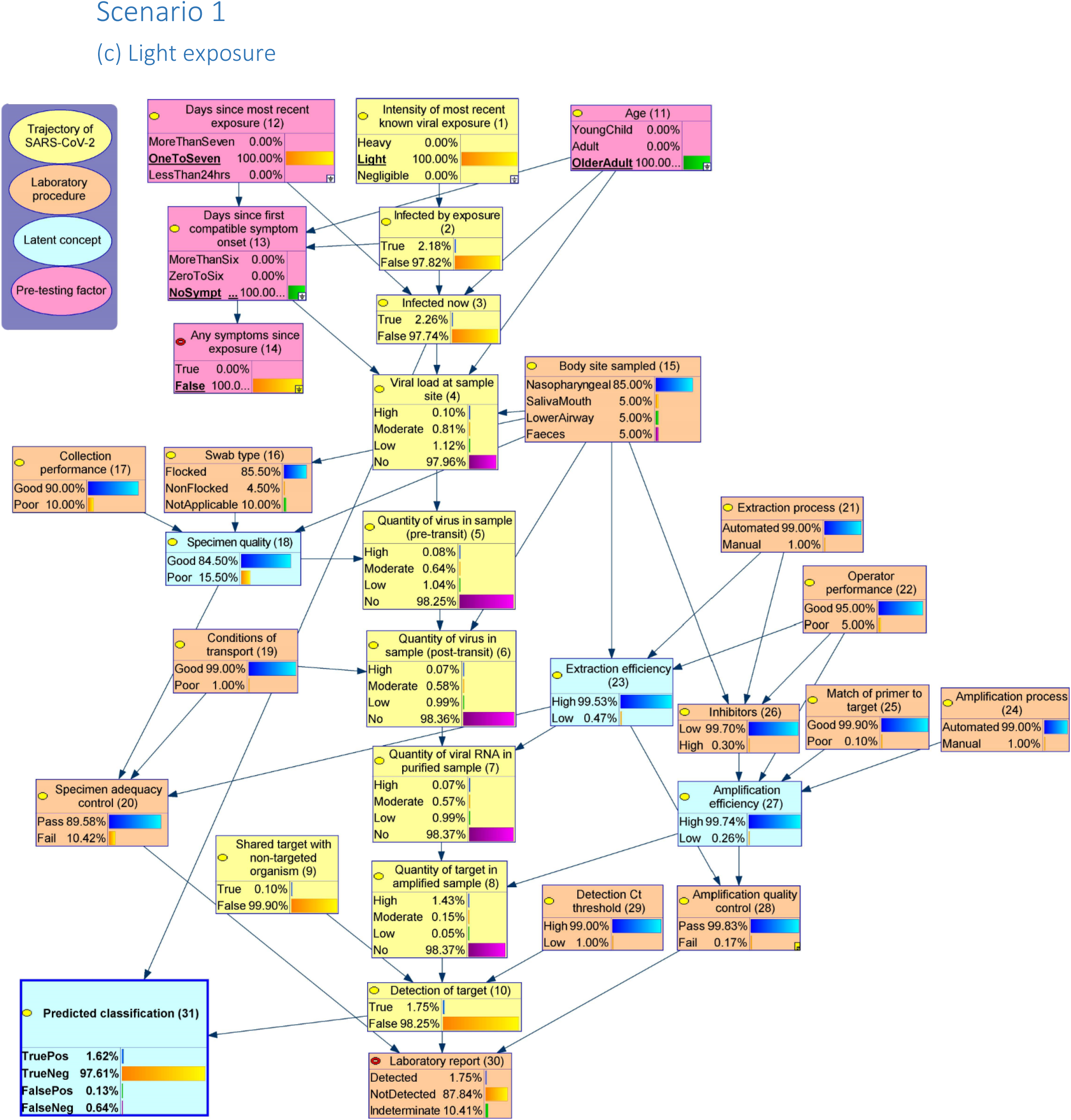
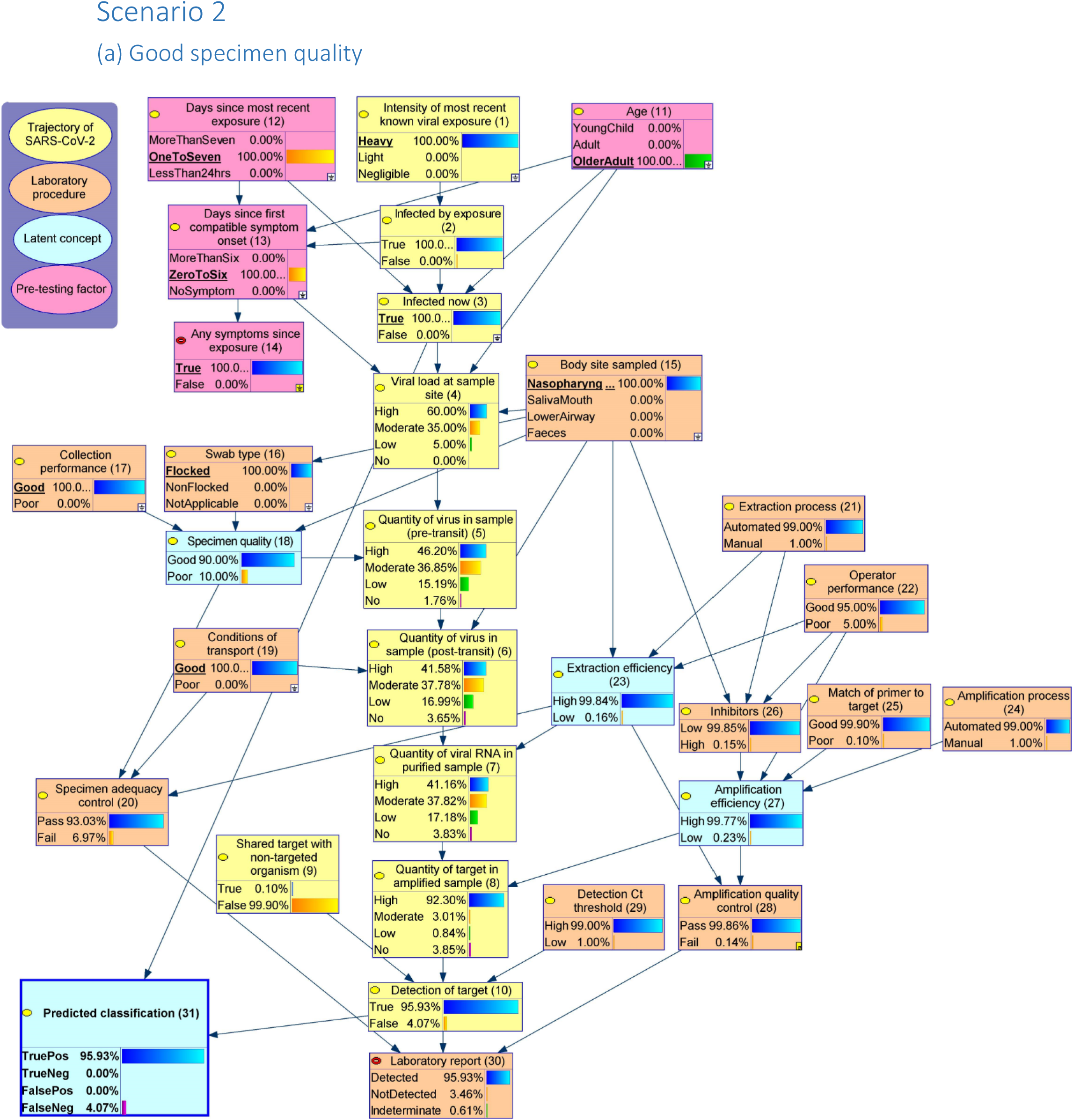
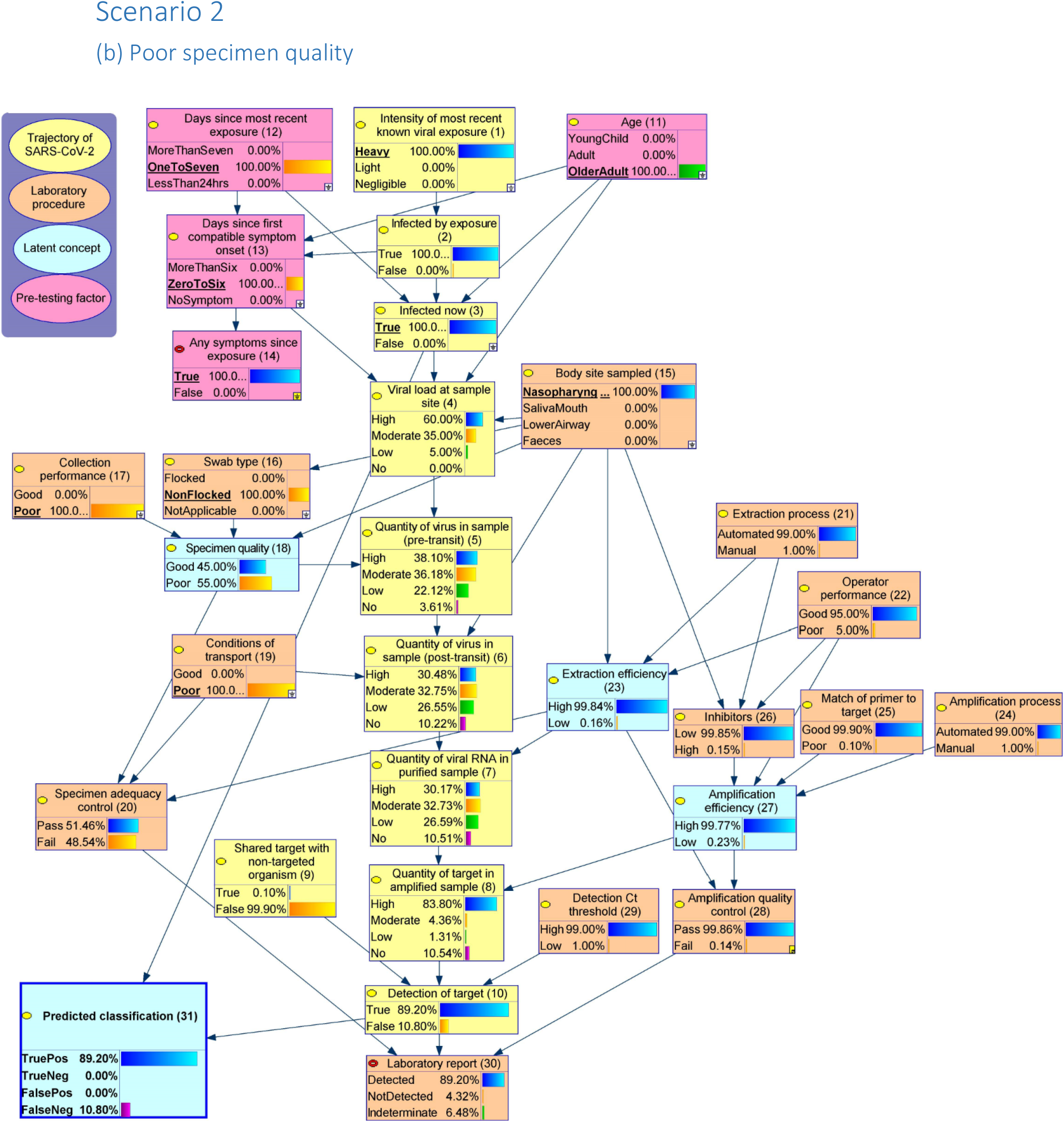
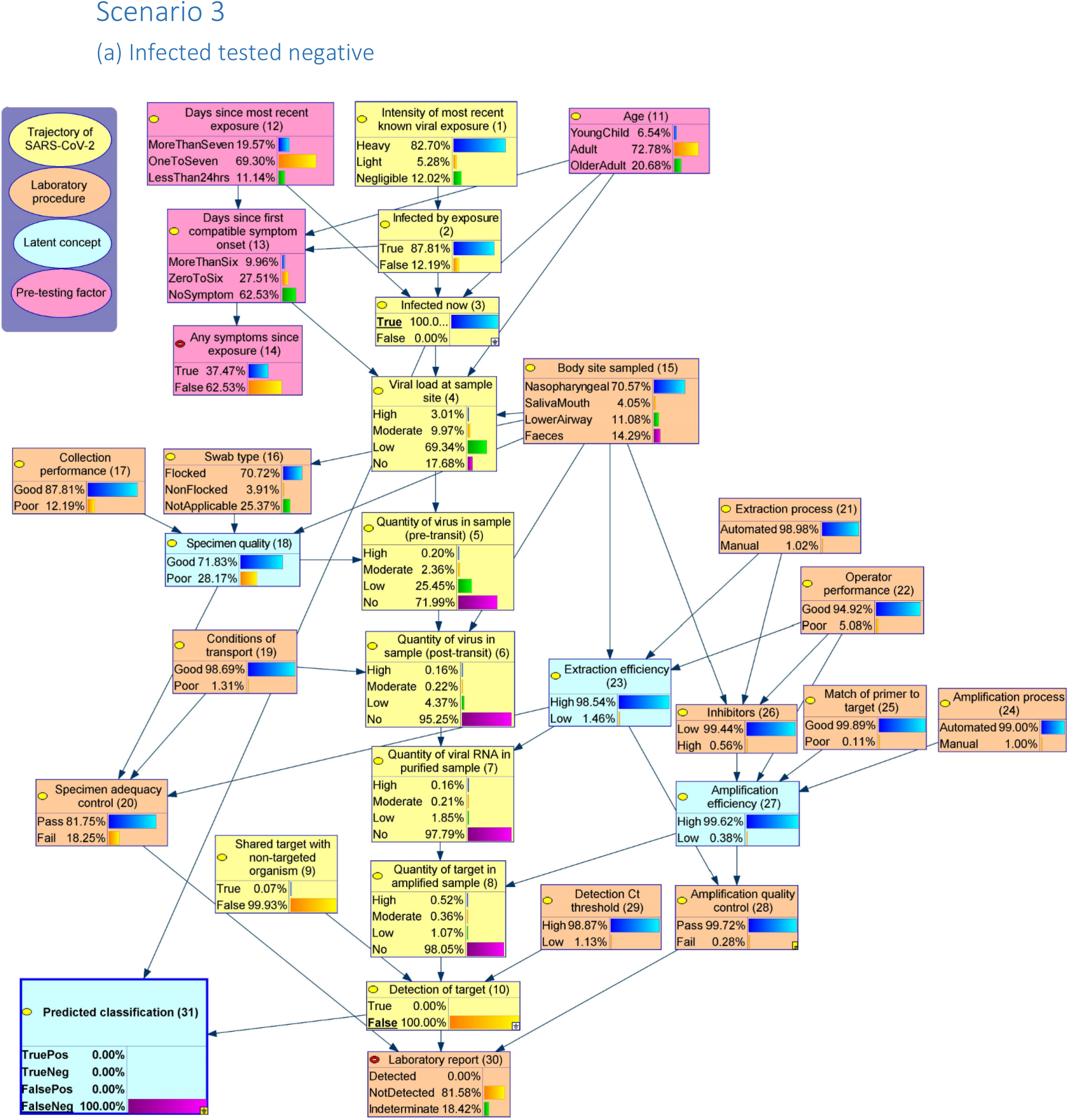
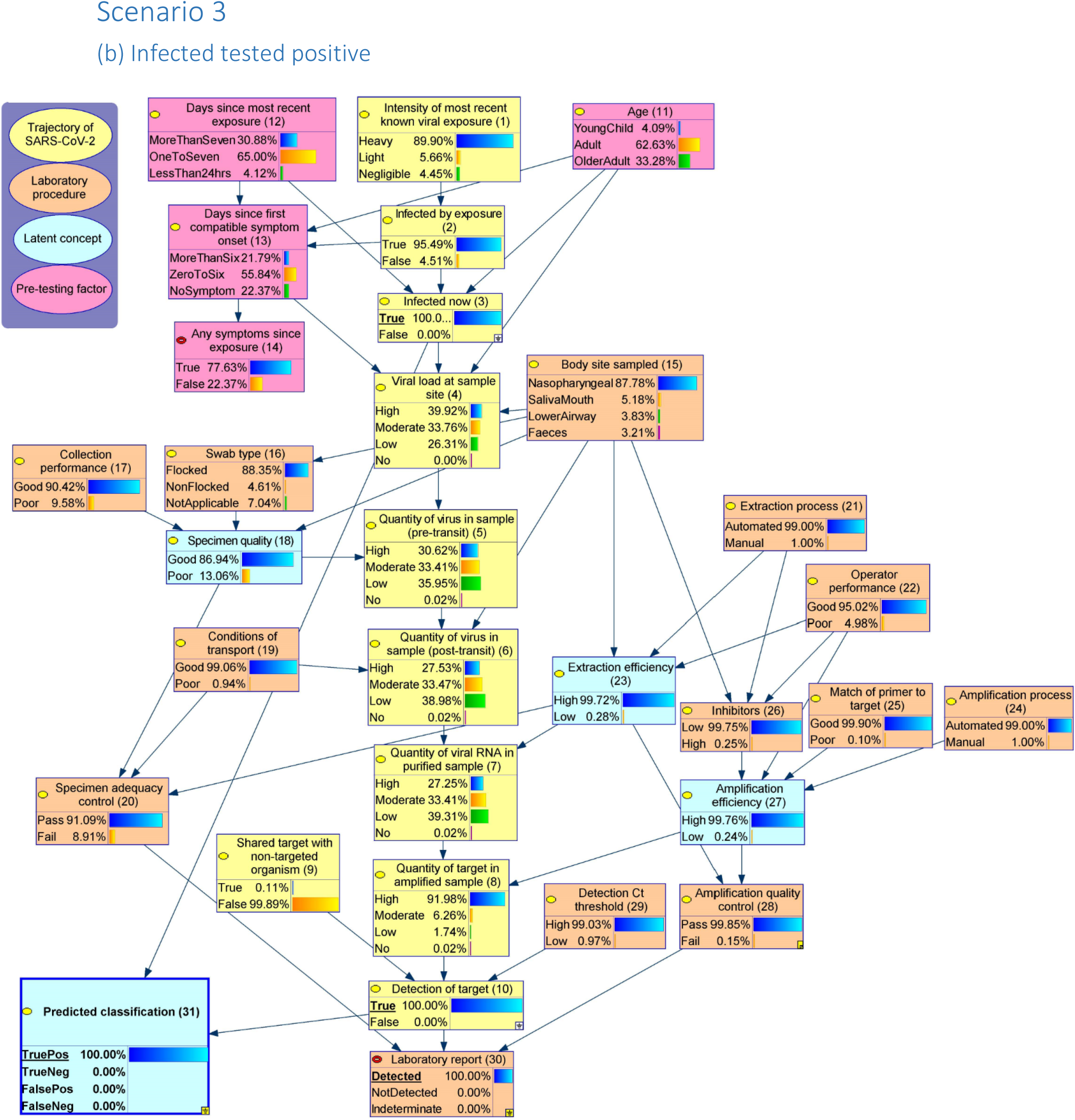
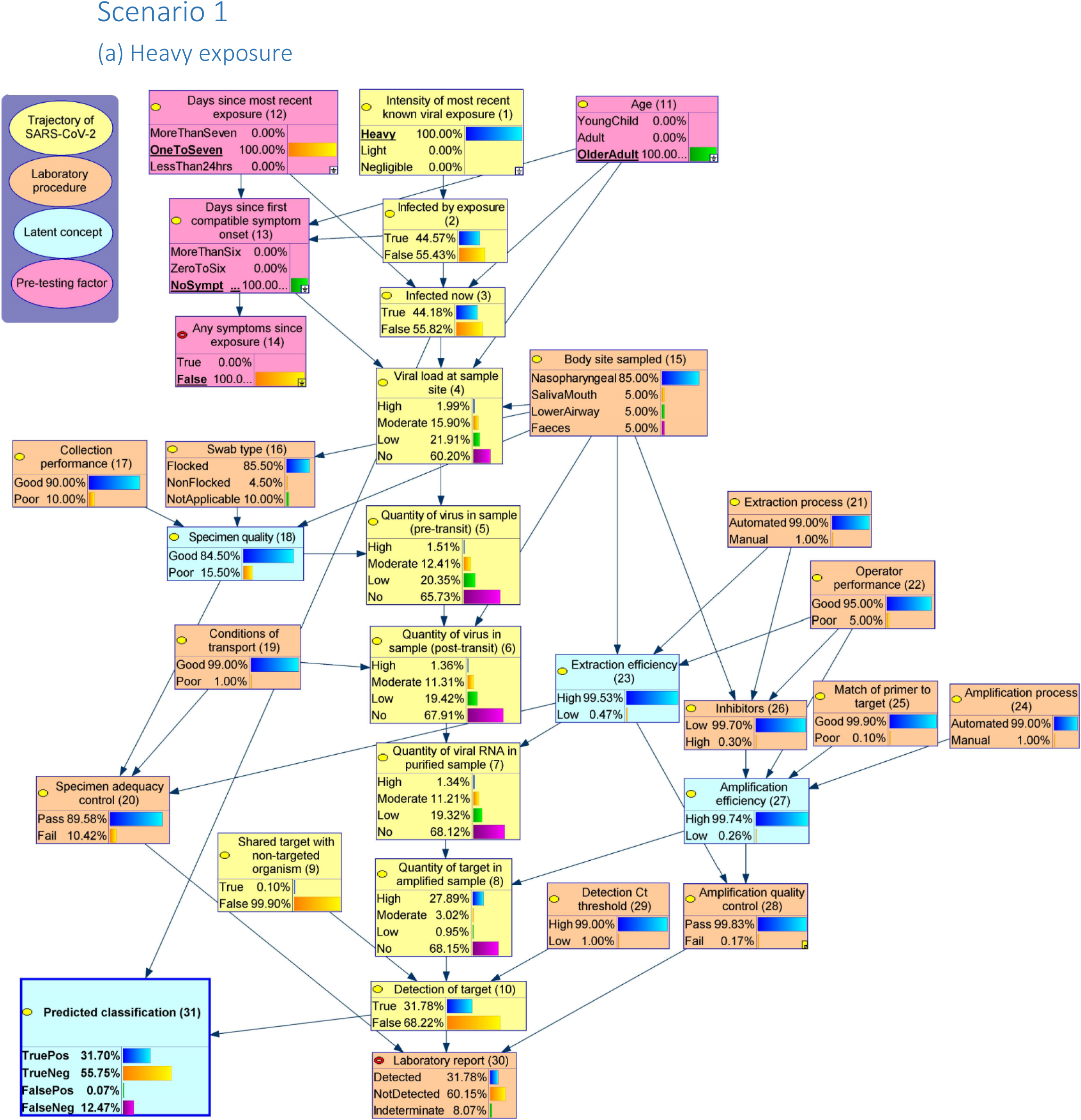
Scenarios.

## Footnotes

i Link to OSF https://osf.io/x5c4u/?view_only=afbdfb3e22c2406cad1ae142a3e5a3b2

ii Node numbers have been kept consistent with the full network in Figure 2.

iii For specific variable definitions, see appendix A, the variable dictionary.

iv The arrow from node (13) to node (4) is the only non-causal link in this model, it summarises events that occur during the time from first symptom onset to testing that may affect viral load at sample site (accumulation or decrease).

## Notes

### Competing Interest Statement

The authors have declared no competing interest.

## References

1. Woloshin S, Patel N, Kesselheim AS. False Negative Tests for SARS-CoV-2 Infection— Challenges and Implications. New England Journal of Medicine. 2020.

2. Arevalo-Rodriguez I, Buitrago-Garcia D, Simancas-Racines D, et al. False-negative results of initial RT-PCR assays for COVID-19: a systematic review. medRxiv. 2020.

3. Kucirka LM, Lauer SA, Laeyendecker O, Boon D, Lessler J. Variation in False-Negative Rate of Reverse Transcriptase Polymerase Chain Reaction-Based SARS-CoV-2 Tests by Time Since Exposure. Ann Intern Med. 2020;173(4):262–267.

4. Pearl J. Embracing causality in default reasoning. Artificial Intelligence. 1988;35(2):259–271.

5. Pearl J. Models, reasoning and inference. Cambridge, UK: CambridgeUniversityPress. 2000.

6. Korb KB, Nicholson AE. Bayesian artificial intelligence. CRC press; 2010.

7. McLachlan S, Dube K, Hitman GA, Fenton N, Kyrimi E. Bayesian Networks in Healthcare: Distribution by Medical Condition. Artificial Intelligence in Medicine. 2020:101912.

8. Marcot BG, Penman TD. Advances in Bayesian network modelling: Integration of modelling technologies. Environmental Modelling & Software. 2019;111:386–393.

9. Larremore DB, Wilder B, Lester E, et al. Test sensitivity is secondary to frequency and turnaround time for COVID-19 surveillance. medRxiv. 2020:2020.2006.2022.20136309.

10. van Kasteren PB, van der Veer B, van den Brink S, et al. Comparison of seven commercial RT-PCR diagnostic kits for COVID-19. Journal of Clinical Virology. 2020;128:104412.

11. Udugama B, Kadhiresan P, Kozlowski HN, et al. Diagnosing COVID-19: The Disease and Tools for Detection. ACS Nano. 2020;14(4):3822–3835.

12. Caruana G, Croxatto A, Coste AT, et al. Diagnostic strategies for SARS-CoV-2 infection and interpretation of microbiological results. Clinical Microbiology and Infection. 2020;26(9):1178–1182.

13. Iwasaki S, Fujisawa S, Nakakubo S, et al. Comparison of SARS-CoV-2 detection in nasopharyngeal swab and saliva. medRxiv. 2020:2020.2005.2013.20100206.

14. Osaro E, Chima N. Challenges of a negative work load and implications on morale, productivity and quality of service delivered in NHS laboratories in England. Asian Pac J Trop Biomed. 2014;4(6):421–429.

15. Stellrecht KA. The Drift in Molecular Testing for Influenza: Mutations Affecting Assay Performance. J Clin Microbiol. 2018;56(3):e01531–01517.

16. Yeoh DK, Foley DA, Minney-Smith CA, et al. The impact of COVID-19 public health measures on detections of influenza and respiratory syncytial virus in children during the 2020 Australian winter. Clinical Infectious Diseases. 2020.

17. Luo L, Liu D, Liao X, et al. Contact Settings and Risk for Transmission in 3410 Close Contacts of Patients With COVID-19 in Guangzhou, China: A Prospective Cohort Study. Annals of internal medicine. 2020;0(0):null.

18. Heald-Sargent T, Muller WJ, Zheng X, Rippe J, Patel AB, Kociolek LK. Age-Related Differences in Nasopharyngeal Severe Acute Respiratory Syndrome Coronavirus 2 (SARS-CoV-2) Levels in Patients With Mild to Moderate Coronavirus Disease 2019 (COVID-19). JAMA Pediatrics. 2020;174(9):902–903.

19. Bi Q, Wu Y, Mei S, et al. Epidemiology and Transmission of COVID-19 in Shenzhen China: Analysis of 391 cases and 1,286 of their close contacts. medRxiv. 2020.

20. Sethuraman N, Jeremiah SS, Ryo A. Interpreting Diagnostic Tests for SARS-CoV-2. JAMA. 2020;323(22):2249–2251.

21. Wiersinga WJ, Rhodes A, Cheng AC, Peacock SJ, Prescott HC. Pathophysiology, Transmission, Diagnosis, and Treatment of Coronavirus Disease 2019 (COVID-19): A Review. JAMA. 2020;324(8):782–793.

22. Kucirka LM, Lauer SA, Laeyendecker O, Boon D, Lessler J. Variation in False-Negative Rate of Reverse Transcriptase Polymerase Chain Reaction–Based SARS-CoV-2 Tests by Time Since Exposure. Annals of internal medicine. 2020;173(4):262–267.

23. Lee S, Kim T, Lee E, et al. Clinical Course and Molecular Viral Shedding Among Asymptomatic and Symptomatic Patients With SARS-CoV-2 Infection in a Community Treatment Center in the Republic of Korea. JAMA Internal Medicine. 2020.

24. Yang Y, Yang M, Shen C, et al. Evaluating the accuracy of different respiratory specimens in the laboratory diagnosis and monitoring the viral shedding of 2019-nCoV infections. medRxiv. 2020:2020.2002.2011.20021493.

25. Azzi L, Carcano G, Gianfagna F, et al. Saliva is a reliable tool to detect SARS-CoV-2. Journal of Infection. 2020.

26. Liu Y, Liao W, Wan L, Xiang T, Zhang W. Correlation Between Relative Nasopharyngeal Virus RNA Load and Lymphocyte Count Disease Severity in Patients with COVID-19. Viral Immunology. 2020;0(0):null.

27. Pujadas E, Chaudhry F, McBride R, et al. SARS-CoV-2 Viral Load Predicts COVID-19 Mortality. medRxiv. 2020.

28. To Kea. Temporal profiles of viral load in posterior oropharyngeal saliva samples and serum antibody responses during infection by SARS-CoV-2: an observational cohort study. The Lancet Infectious Diseases. 2020;20(5):565–574.

29. Pan Y, Zhang D, Yang P, Poon LL, Wang Q. Viral load of SARS-CoV-2 in clinical samples. The Lancet Infectious Diseases. 2020;20(4):411–412.

30. Garnett L, Bello A, Tran KN, et al. Comparison analysis of different swabs and transport mediums suitable for SARS-CoV-2 testing following shortages. J Virol Methods. 2020;285:113947–113947.

31. Centers for Disease Control Prevention. Interim guidelines for collecting, handling, and testing clinical specimens from persons under investigation (PUIs) for coronavirus disease 2019 (COVID-19). COVID-19 Web site. Published 2020. Updated 8th July 2020. Accessed 22nd September, 2020.

32. McCulloch DJ, Kim AE, Wilcox NC, et al. Comparison of Unsupervised Home Self-collected Midnasal Swabs With Clinician-Collected Nasopharyngeal Swabs for Detection of SARS-CoV-2 Infection. JAMA Network Open. 2020;3(7):e2016382–e2016382.

33. Tu Y-P, Jennings R, Hart B, et al. Swabs Collected by Patients or Health Care Workers for SARS-CoV-2 Testing. New England Journal of Medicine. 2020;383(5):494–496.

34. Fang Y, Zhang H, Xie J, et al. Sensitivity of Chest CT for COVID-19: Comparison to RT-PCR. Radiology. 2020;296(2):E115–E117.

35. Raschke RA, Curry SC, Glenn T, Gutierrez F, Iyengar S. A Bayesian Analysis of Strategies to Rule Out Coronavirus Disease 2019 (COVID-19) Using Reverse Transcriptase–Polymerase Chain Reaction. Archives of Pathology & Laboratory Medicine. 2020;144(8):915–916.

36. Wang W, Xu Y, Gao R, et al. Detection of SARS-CoV-2 in Different Types of Clinical Specimens. JAMA. 2020;323(18):1843–1844.

37. Young BE, Ong S, Kalimuddin S, et al. Epidemiologic Features and Clinical Course of Patients Infected With SARS-CoV-2 in Singapore. JAMA. 2020;323(15):1488–1494.

38. Wyllie AL, Fournier J, Casanovas-Massana A, et al. Saliva or Nasopharyngeal Swab Specimens for Detection of SARS-CoV-2. New England Journal of Medicine. 2020.

39. Ali N, Rampazzo RdCP, Costa ADT, Krieger MA. Current Nucleic Acid Extraction Methods and Their Implications to Point-of-Care Diagnosticsvol. BioMed Research International. 2017;2017:13 pages.

40. Hammerling JA. A Review of Medical Errors in Laboratory Diagnostics and Where We Are Today. Laboratory Medicine. 2012;43(2):41–44.

41. Broeders S, Huber I, Grohmann L, et al. Guidelines for validation of qualitative real-time PCR methods. Trends in Food Science & Technology. 2014;37(2):115–126.

42. Sung H, Han M-G, Yoo C-K, et al. Nationwide External Quality Assessment of SARS-CoV-2 Molecular Testing, South Korea. Emerging Infectious Diseases. 2020;26(10).

43. Schrader C, Schielke A, Ellerbroek L, Johne R. PCR inhibitors – occurrence, properties and removal. Journal of Applied Microbiology. 2012;113(5):1014–1026.

44. Brukner I, Eintracht S, Papadakis AI, et al. Maximizing confidence in a negative result: Quantitative sample adequacy control. Journal of Infection and Public Health. 2020;13(7):991–993.

45. Vogels CBF, Brito AF, Wyllie AL, et al. Analytical sensitivity and efficiency comparisons of SARS-COV-2 qRT-PCR primer-probe sets. medRxiv. 2020:2020.2003.2030.20048108.

46. Nalla AK, Casto AM, Huang M-LW, et al. Comparative Performance of SARS-CoV-2 Detection Assays Using Seven Different Primer-Probe Sets and One Assay Kit. Journal of Clinical Microbiology. 2020;58(6):e00557–00520.

47. Corman VM, Landt O, Kaiser M, et al. Detection of 2019 novel coronavirus (2019-nCoV) by real-time RT-PCR. Eurosurveillance. 2020;25(3):2000045.

48. Osório NS, Correia-Neves M. Implication of SARS-CoV-2 evolution in the sensitivity of RT-qPCR diagnostic assays. The Lancet Infectious Diseases. 2020.

49. Han MS, Byun J-H, Cho Y, Rim JH. RT-PCR for SARS-CoV-2: quantitative versus qualitative. The Lancet Infectious Diseases. 2020.

50. Espy MJ, Uhl JR, Sloan LM, et al. Real-time PCR in clinical microbiology: applications for routine laboratory testing. Clinical microbiology reviews. 2006;19(1):165–256.

51. Woloshin S, Patel N, Kesselheim AS. False Negative Tests for SARS-CoV-2 Infection — Challenges and Implications. New England Journal of Medicine. 2020;383(6):e38.

52. Watson J, Whiting PF, Brush JE. Interpreting a covid-19 test result. Bmj. 2020;369:m1808.

